# Identifying genetic variants causal for multiple long-term conditions: informing opportunities for intervention and prevention

**DOI:** 10.64898/2026.09.28.26364012

**Authors:** Bethany Voller, Ruby M Woodward, Olivia Murrin, João Delgado, Deniz Türkmen, Rhian Hopkins, Concepción Violán, Mary Mancini, Leon Farmer, Kate Boddy, Chris Fox, Sarah E Lamb, Frank Dudbridge, Jack Bowden, Timothy M Frayling, Jane AH Masoli, Luke C Pilling, the GEMINI Consortium

## Abstract

Multiple long-term conditions (MLTC) – the coexistence of two or more long-term conditions (LTCs) in an individual, is an increasingly prevalent and complex clinical challenge. We aimed to uncover opportunities for therapeutic intervention in MLTC and highlight potential adverse effects using genetics to identify shared biological pathways across conditions.

Across 72 heritable LTCs common in adults aged ≥65, we analysed genome-wide association study (GWAS) summary statistics from meta-analyses incorporating up to three data sources (UK Biobank, FinnGen, and condition-specific consortia) from genetically European-ancestry participants. Across 843 condition pairs sharing genome-wide significant variants, we performed statistical colocalisation to identify shared causal genetic variants. Variants showing evidence of causality for ≥3 LTCs were prioritised for investigation of likely causal genes, biological pathways, and druggability. Findings were replicated in Our Future Health (OFH, N=550,000).

Colocalisation analyses identified 281 genetic variants causal for specific LTC pairs and 85 variants causal for ≥3 LTCs. These 85 variants were located in 34 genomic regions with strong evidence of shared causality across multiple conditions. Thirteen of the 34 regions fully replicated in OFH, with others having partial replication. Eight variants showed concordant directions of effects across conditions, suggesting potential opportunities for drug development; results include a variant in *GDF7* causal for diverticular disease, gastro-oesophageal reflux disease, and prostate cancer, suggesting multisystemic mechanisms and a novel intervention target for this cluster of conditions. We identified six potential drug repurposing opportunities involving clinically targeted proteins, including LPA and IRF5 for cardiovascular and immune-mediated inflammatory conditions, respectively. Fifteen variants showed discordant effects across phenotypes, suggesting possible adverse consequences of pharmacological intervention; for example, a variant in *GIPR* reduced risk of obesity and sleep apnoea whilst increasing risk of breast cancer.

These findings reveal shared mechanisms underlying MLTC and highlight opportunities for therapeutic development and repurposing, whilst also identifying possible adverse effects.

**Author summary:** Many individuals live with multiple long-term health conditions, but relatively little is known about the biological processes linking these conditions together. Where conditions share biology, this could reveal new ways to treat diseases. In this study, we used data from large volunteer cohorts of hundreds of thousands of individuals to find regions of DNA associated with multiple conditions. DNA codes for genes, and we found specific genes and biological pathways that partially explain why some conditions occur together. For example, we found evidence linking diverticular disease, gastro-oesophageal reflux disease, and prostate cancer, suggesting that these seemingly different conditions may share underlying biological mechanisms. We identified several proteins that could be explored as new drug targets and opportunities to repurpose existing medications. We also found evidence suggesting that some treatments could have unintended effects, reducing the risk of one condition but increasing the risk of another. This work may help support the development of treatments for people with multiple long-term conditions.

## Introduction

The coexistence of multiple long-term health conditions (MLTC) in an individual, also referred to as ‘multimorbidity’, is a complex clinical problem affecting over 50% of adults aged 65 and over [1]. Clinical guidelines, health service and research delivery are largely focused on the treatment and prevention of single diseases; yet this may not be optimum for individuals with MLTC. Individuals with MLTC are often prescribed multiple different medicines (polypharmacy) which can interact with each other, potentially leading to adverse drug reactions [2]. This can increase the burden on both the individual and the health care system. Despite this, individuals with MLTC are underrepresented in clinical trials relative to comparable real-world populations, limiting the applicability of trial evidence to people with MLTC [3]. Knowledge is therefore needed of which conditions co-occur more often than would be expected by chance, and the possible mechanisms driving these associations. The GEMINI (Genetic Evaluation of Multimorbidity towards INdividualisation of Interventions) collaborative curated and defined a set of 72 LTCs that are common in people aged ≥65 and had significant genetic heritability [4]. We conducted pairwise observational and genetic analyses of the 72 LTCs, showing the breadth of LTC co-occurrence and the extent of shared genetic mechanisms.

Genetic studies are key to understanding the mechanisms underlying observed condition co-occurrence and identifying potential therapeutic targets; drug targets with human genetic evidence are more than twice as likely to be approved [5,6]. Genome-wide association studies (GWAS) have identified many genetic variants influencing individual diseases; investigating variants that have causal effects on numerous distinct phenotypes – pleiotropy – can improve understanding of shared biological mechanisms and inform pharmacological interventions. A 2019 study of pleiotropy across 558 traits found widespread evidence of shared variants and genes associated with more than one trait [7]. Further, genetic correlation analyses have revealed extensive shared genetic influences in MLTC, suggesting overlapping causal mechanisms [4,8,9]. However, genetic correlations represent an average measure of how genetic effects align across the genome. Colocalisation methods can identify the individual shared causal variants between traits [10], revealing shared aetiology and underlying biological pathways between them [10–12].

Integrative GWAS and colocalisation analysis using publicly available genetic resources such as the Open Targets Platform [13] can identify genetic signals shared between two or more traits and assess their potential as drug targets. A recent study included 30 age-related diseases and highlighted the potential of using genetic approaches to identify opportunities for drug development in MLTC [14]. Whilst previous studies have explored shared genetic architecture in MLTC, and linked findings to opportunities for drug development, studies interrogating directionality across MLTC to highlight potential risks of adverse treatment effects where there are discordant directions of effect are limited; though a recent Open Targets study of pleiotropy in GWAS reports directional consistency across many identified loci [15].

In this study, we use genetic colocalisation to search for shared causal mechanisms between 2,211 pairs of LTCs. By investigating the biological pathways involved, we explore whether these variants could be promising targets for novel interventions or repurposing opportunities in MLTC or whether they highlight potential risks of adverse interactions.

## Methods

### Genetic data and sources

We used genome-wide association study (GWAS) summary statistics from the GEMINI (Genetic Evaluation of Multimorbidity towards INdividualisation of Interventions) collaborative. The first GEMINI study ascertained 72 common and heritable long-term conditions [4], and for each condition, meta-analysed GWAS data from up to three sources: two large cohort studies, the UK Biobank (N=502,000)[16] and FinnGen (release 9; N=377,000) [17], and condition-specific consortium data when available. Full details are described in the study [4], and curated diagnostic code lists for all 72 conditions can be found on the GEMINI GitHub (https://github.com/GEMINI-multimorbidity). Analyses only included participants genetically similar to the 1000 Genomes European subset, due to the requirements of software used and data available.

Linkage disequilibrium data (R^2^ and D′) reported throughout the manuscript are from LDlink EUR population [18].

### Exploring shared genetic variants and loci across MLTC

For all 72 LTCs, we identified the genome-wide significant variants (p<5×10^-8^). We defined distance-based genetic loci for each condition: starting with the variant with the smallest p-value, we defined regions of ±250kb around each variant; this was done iteratively until all variants had been assigned a region. The number of variants reaching genome-wide significance and the number of genetic loci is described in Supplementary Table 1. To explore shared genetic architecture across conditions, we defined a unified set of global loci. First, physically overlapping loci within individual conditions were merged, and then overlapping loci across conditions were merged. Within a single global locus, not all individual condition-associated loci necessarily overlapped; if locus A overlapped with locus B, and locus B overlapped with locus C, but loci A and C did not overlap, they would still be in the same global locus (see Supplementary Figure 1 for illustration).

### Identifying shared causal mechanisms using genetic colocalisation

For all pairwise combinations of LTCs, we identified shared genome-wide significant variants, and starting with the variant with the smallest p-value, defined regions of ±250kb iteratively until all variants had been assigned a region. To formally test whether these regions contained variants causal for both LTCs, we performed statistical colocalisation using the *coloc.abf* function from the coloc R-package (version 5.2.3) [10]. The *coloc.abf* function assumes a single causal variant for each trait in a given genomic region.

Coloc is a Bayesian method that calculates the posterior probabilities of five hypotheses for each given genomic region for a pair of traits. As described in our prior study [19], we focused on hypotheses 3 and 4 (H3 and H4): H3 is the hypothesis that there are two distinct causal variants in a region, one for each trait; H4 is the hypothesis that there is a single causal variant in the region, common to both traits [10]. We used the default prior probabilities: p_1_=1×10^-4^, p_2_=1×10^-4^, p_12_=1×10^-5^, where p_1_ and p_2_ are the probabilities that a random SNP in the region is causally associated with trait 1 or trait 2 respectively, and p_12_ is the probability that a variant is causal for both traits. The default value p_12_=1×10^-5^ is more likely to detect false shared associations than a more conservative smaller choice of p_12_ but may also lead to missing valid shared causal variants with slightly weaker signals [20].

To conclude sufficient evidence for colocalisation in any given genomic region, we looked for a high combined posterior probability of H3 and H4 (PPH3 and PPH4, respectively) as this suggests an association with both traits (regardless of whether the exact cause is the same for both traits): PPH3+PPH4≥0.8, and if PPH4/PPH3>5, as this suggests sufficient support to consider that it is a ‘convincing’ colocalising signal [11].

For the LTC pairs with evidence of colocalisation in at least one genetic region, we extracted the posterior probabilities for each variant in the region to be causal conditional on H4 being true. These variants were filtered to those with an individual probability of being the specific shared causal variant greater than 50%. We then identified the variants that appeared for more than one LTC pair (i.e. three or more individuals LTCs) and grouped them into regions based on distance – if a variant was more than 1Mb from the previous one, it was classified as a new region. We removed variants in and around the HLA region on chromosome 6 (GRCh37 position 28.5Mb-33.5Mb), as this region is known to have exceptionally complex patterns of linkage disequilibrium (LD) that could confound analyses. We chose to prioritise genetic variants that were implicated across three or more LTCs as variants associated with multiple conditions may represent more highly pleiotropic genetic signals, which could have more relevance in MLTC.

### Mechanistic consequences of identified causal variants

For the variants with high probability of being causal for multiple LTC pairs, we used function ‘query_GWAS_Catalog()’ from R package {timesaver} (v0.0.1.6 https://github.com/alesssia/timesaveR) to search the European Bioinformatics Institute (EBI) GWAS Catalog (https://www.ebi.ac.uk/gwas/) [21] for traits previously associated with each variant, along with traits associated with variants in strong LD (R^2^>0.9). We used the University of California Santa Cruz (UCSC) Genome Browser [22] to explore the genomic position of each variant, and the Open Targets Platform [13] (https://platform.opentargets.org/) to determine the nearest gene. If a variant did not lie within a gene, the nearest gene was defined as the nearest protein-coding gene with the closest transcription start site.

To assist in linking variants to genes, we looked to see whether variants were associated with levels of circulating proteins – whether they were protein quantitative trait loci (pQTLs). We used data on 2,923 proteins from the UK Biobank Pharma Proteomics Project (N=48,195 European individuals), profiled using Olink technology [23]. We used a suggestive GWAS significance threshold of p<5×10^-5^ to identify significant pQTL-LTC associations. We classified *cis*-pQTLs as those residing within 944kb of the transcription start site for the gene encoding the protein [24]; the rest were defined as *trans*-pQTLs. We also used the Genotype-Tissue Expression database v10 (GTEx, https://gtexportal.org/home/) [25] to explore whether variants were associated with the expression levels of genes – whether they were expression quantitative trait loci (eQTLs). We annotated variants using an extension of the Open Targets Platform, the Enhancer-Gene (E2G) Portal (https://e2g.stanford.edu/), which maps variants to target genes based on genomic overlap with predicted enhancer regions.

### Exploring the druggability of linked proteins

To investigate whether we could identify any targets for prevention or intervention in MLTC, we explored the *druggability* of proteins encoded by the genes linked to variants. ‘Druggability’ describes the ability of a protein to interact with drug-like molecules [26]. We used the Druggable Genome database [26] along with the Open Targets Platform – specifically the tractability data [13] to identify proteins with potential to be drug targets. Tractability assessments are generated for small molecule and antibody modalities, along with proteolysis targeting chimeras (PROTACs). PROTACs are an emerging drug modality designed to destroy disease-causing proteins, rather than just blocking their activity, by hijacking the ubiquitin-proteasome system of cells (the cell’s protein recycling system), tagging disease-causing proteins for degradation [27]. In our study, we classify a protein as druggable if it is considered druggable in the druggable genome database, or if there is tractability evidence from Open Targets.

For druggable proteins, we defined whether they were clinically targeted using two levels of therapeutic evidence: target-level and pathway-level. We classified a protein as clinically targeted with target-level therapeutic evidence if there was an existing intervention, either approved or in clinical trials, directly modulating the protein’s function, representing direct pharmacological manipulation of the protein. Therapeutic interventions were classified as pathway-level evidence if they were approved or in clinical trials (see below), and do not directly interact with the protein itself, but produce a biologically plausible effect on the same pathway in which the protein acts, indirectly modulating its activity. We did not include interventions without a clear mechanistic link to the implicated protein – e.g. general anti-inflammatory drugs.

Therapeutic evidence was identified using the publicly available databases DrugBank [28] and the Therapeutic Target Database [29], alongside targeted literature searches in PubMed, supplemented by targeted web-based searches. Searches used combinations of gene/protein names with terms related to therapeutic targeting (e.g. ‘drug’, ‘therapy’, ‘therapeutic’, ‘clinical trial’, and ‘approved’). Protein names were queried in ClinicalTrials.gov (https://clinicaltrials.gov/) to explore human trial evidence. Findings were manually reviewed to confirm mechanistic relevance to the implicated protein or pathway.

### Replication in Our Future Health

We used Our Future Health (OFH) release 13 (N=550,000 with imputed genotypes and linked hospital diagnosis records) to replicate variant associations with LTCs (approved study ID OFHS250013). LTCs were ascertained using the same clinical codes lists used throughout GEMINI (https://doi.org/10.5281/zenodo.14824760). For the genetic variants with high probability of being causal for three or more LTCs, we ran logistic regression models to test the variant-trait associations in OFH, adjusted for age, sex, assessment region, genetic principal components (PCs) 1-10, in participants who self-reported any ‘white’ ethnicity (genetic ancestry data was not available at the time of analysis). Final sample size was 472,811. We highlight the regions from our colocalisation analysis for which the specific genetic variants were significantly associated (FDR-adjusted per variant) with the same LTCs in OFH.

## Results

We identified 13 genomic regions containing variants with high probability for shared causality in at least two condition pairs (3 or more distinct diagnoses) that are considered ‘druggable’ and had consistent associations in Our Future Health. Two are already therapeutically targeted (or an intervention is in clinical trials) and may present repurposing opportunities, two are already targeted but discordant effect directions may indicate adverse treatment effects, and five are potentially novel therapeutic targets. See Figure 1 for analysis flowchart and Figure 3 for variant-disease-protein effects. Results are described in detail below.

**Figure 1:**
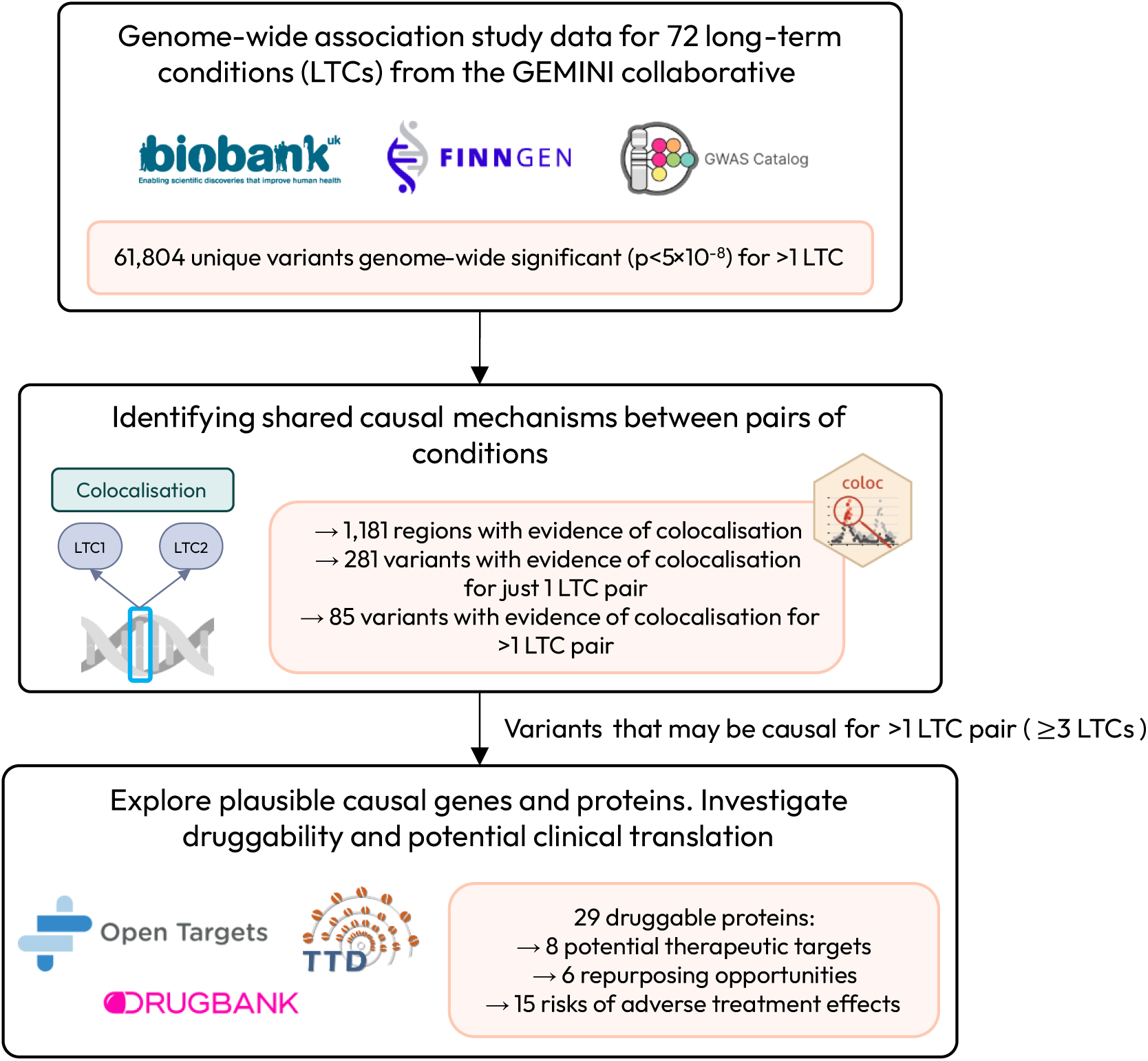
Analysis flowchart. Flowchart illustrating the steps of the genetic analyses performed, along with key results.

### Shared genetic loci across 72 LTCs

We tested the extent of shared genetic architecture across conditions. Across all 72 LTCs, 267,778 variants were genome-wide significant for at least one condition (Supplementary Figure 2; Supplementary Table 2). Of these, 61,804 (23.1%) were associated with more than one condition, of which 28,430 (10.6%) were associated with three or more conditions (Supplementary Table 2, Supplementary Figure 2). Looking at pairwise combinations across the 67 LTCs with genome-wide significant (p<5×10^-8^) variants (five conditions had none: enthesopathies of the lower body, essential tremor, fibromyalgia, insomnia, and peripheral neuropathies), 843 condition pairs (out of a possible 2,211) shared genome-wide significant variants. We mapped these to 1,324 loci across the 67 LTCs, indicating widespread shared genetic architecture across the conditions. The number of loci per condition is shown in Supplementary Figure 4, and an illustration of the global loci across the 67 LTCs can be seen in Supplementary Figure 5.

### Shared causal mechanisms identified using colocalisation

To identify which conditions shared causal genetic variants we performed colocalisation analyses. For a pair of traits, colocalisation allows us to assess the probability that given genomic regions contain a genetic variant that is causal for both conditions. We did this for condition pairs that shared genome-wide significant variants – 843 condition pairs, first looking at shared causal loci, and then at specific shared causal variants. Figure 2 is a flowchart illustrating the colocalisation findings.

**Figure 2:**
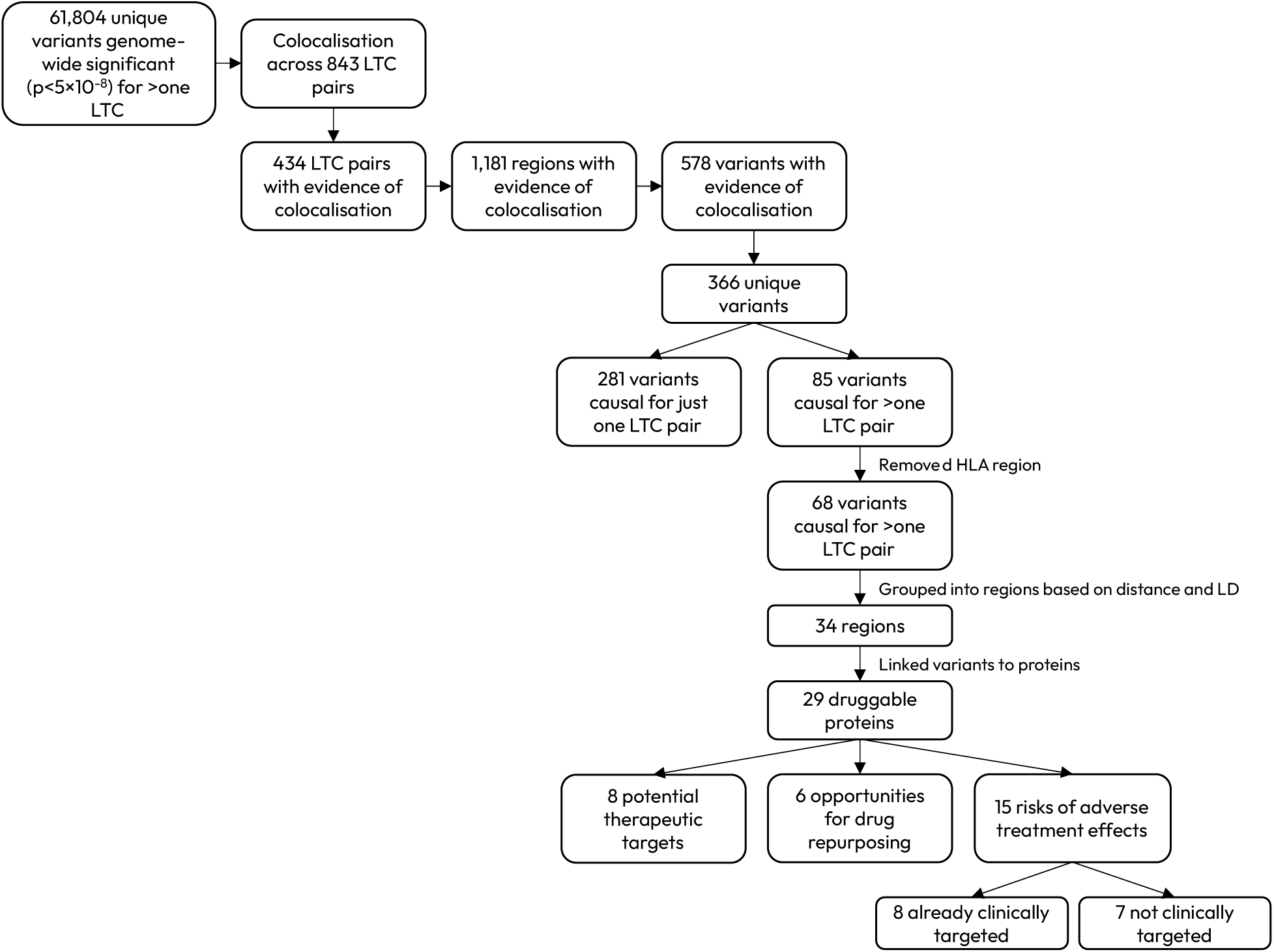
Flowchart illustrating colocalisation process. A flowchart illustrating each step of the analysis process, from genome-wide significant variants to identifying potential therapeutic targets, repurposing opportunities, and risks of adverse treatment effects.

Colocalisation analysis identified 434 LTC pairs with one or more colocalising regions: regions with high probability of a single genetic variant causal for both conditions (Supplementary Table 3). There was a total of 1,181 regions with evidence of colocalisation across the 434 pairs; the condition pair sharing the most colocalising regions was coronary heart disease and hypertension, both cardiovascular conditions (40 colocalising regions), whilst a cross-domain condition pair, hypertension and type 2 diabetes, shared 33 colocalising regions, the second highest number. We identified 366 specific variants with evidence of causing at least one LTC pair. Of these, 85 variants were causal for more than one condition pair; after removing those in the HLA region, 68 variants remained. We grouped these into 34 regions based on position on the genome and LD (Supplementary Table 4).

We classified regions into different groups, depending on whether the variant(s) in the region showed concordant or discordant directions of effect on all colocalising conditions in the region, whether the implicated protein has potential to be druggable or not, and whether the protein is already targeted (with either target-level or pathway-level evidence as previously defined) by approved drugs or drugs in clinical trials. For 29 regions, the variants linked to proteins that are either already drugged by approved drugs or drugs in clinical trials or have potential to be drugged. These regions are illustrated in Figure 3, which shows for each region: the LTCs that the variant was found to be causal for, the gene that we linked each variant to, the level of evidence supporting the link, and the potential clinical implication. In cases where a region contained more than one potential causal variant, we selected one variant to show on the figure for visualisation. These regions are indicated on the figure by an asterisk next to the variant ID, and all variants can be found in Supplementary Table 4.

**Figure 3:**
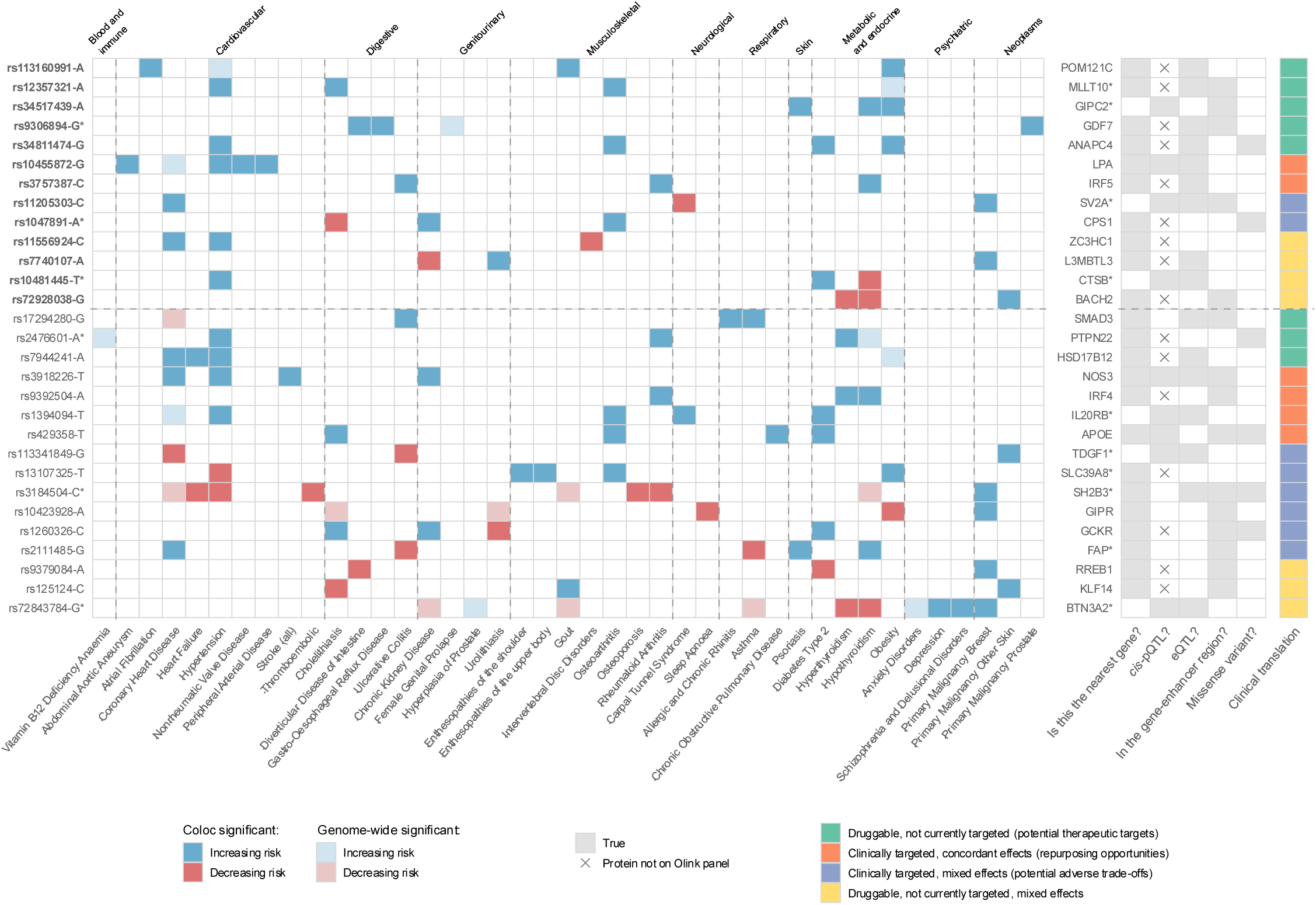
Genetic variants with evidence of causing ≥3 long-term conditions and evidence of druggability, annotated with likely causal protein and clinical translation. Results are from colocalisation analysis of GEMINI long-term conditions. The variants identified with high probability of causing multiple LTCs are shown with direction of effect. The allele shown for each condition was selected to show the cancer risk-increasing allele, or if there was no cancer association, the allele associated with the highest number of LTCs. Variants can be associated with other LTCs (p<5×10^-8^) but not be significant in colocalisation: these are shown in lighter shades of red/blue. White squares indicate p>5×10^-8^ for that variant-LTC association (of all possible variant-LTC associations shown in this heatmap, there were two cases where a variant was not present in the GWAS summary statistics of an LTC). The horizontal dashed line indicates that all variant-LTC associations above the line replicated in Our Future Health with FDR-p<0.05. The vertical dashed lines separate the LTCs by ICD disease domains. An asterisk following a variant ID indicates multiple potential causal variants in that region. An asterisk following a protein name indicates multiple potential proteins in the causal pathway. See Supplementary Tables 4 and 6 for details.

### Replication in Our Future Health

Four of the 68 causal variants were not in the imputed genetic data from OFH (rs149866169, rs28796334, rs7953257, rs369234167). We used LDlink data and identified proxies in OFH (GBR reference panel, R^2^>0.8) for two of the variants (rs7953257=rs17630235 [R^2^=0.81], rs369234167=rs17662749 [R^2^=0.91]). Of the remaining 64 variants (plus 2 proxies), 22 variants in 16 colocalising regions showed significant effects in consistent directions on the same LTCs identified in our primary analysis. These variants are indicated in Supplementary Table 5, and are the ones focused on in the rest of this manuscript. The regions with OFH replication are also highlighted in Figure 3; it should be noted that only 13 regions are shown as replicating in the figure – this is because two of the 16 proteins corresponding to a replicated variant are not considered druggable, so are not shown in the figure. For the remaining region, the region contained multiple potential causal variants; we selected a missense variant (rs3184504) to show in the figure which did not replicate in OFH, though four other variants in the same region did.

### Identifying potential therapeutic targets

We first explored proteins that could be potential novel therapeutic targets. We looked at regions in which the colocalising variant(s) showed concordant directions of effect on all conditions, with the implicated protein thought to be druggable but not currently targeted by any approved drugs or drugs in clinical trials. Colocalisation identified variants which we grouped into eight regions, mapped to the genes *GIPC2*, *GDF7*, *ANAPC4*, *POM121C*, *MLLT10*, *PTPN22*, *SMAD3*, and *HSD17B12*. For five regions (implicating genes *GIPC2*, *POM121C*, *MLLT10*, *GDF7*, and *ANAPC4*), the variants highlighted by colocalisation analysis showed significant effects in consistent directions on the same LTCs in OFH. Details of these five regions, including the colocalising variants, the conditions that they are causal for, and evidence linking variants to genes and proteins, are in Supplementary Tables 4 and 6. The variant linked to *GIPC2* (rs34517439), implicated in hypothyroidism, psoriasis, and obesity, lies in the intron of *DNAJB4* but is a *cis*-pQTL for *GIPC2*, and is thought to overlap with the enhancer region for *GIPC2*, suggesting *GIPC2* may be the more likely effector gene.

For the remaining three regions, two contained clusters of conditions from the same domain; a variant (rs7944241) upstream of *HSD17B12* was causal for a cluster of cardiovascular conditions: coronary artery disease (CAD), heart failure, and hypertension, and a variant (rs1729280) in the intron of *SMAD3* showed causal effects for a group of inflammatory conditions: asthma, allergic and chronic rhinitis, and ulcerative colitis (UC). The third region contained two variants with evidence of causing conditions across different disease domains: vitamin B12 deficiency, hypertension, hypothyroidism, and rheumatoid arthritis (RA). The variants, rs6679677 and rs2476601, upstream of *PHTF1* and a missense variant in *PTPN22* respectively, are in near-complete LD (R^2^=0.97, D′=1).

### Identifying opportunities for drug repurposing

We explored potential opportunities for drug repurposing, where an existing drug could be used in the treatment of additional conditions. We looked at regions in which the colocalising variant(s) showed concordant directions of effect on all conditions, and the implicated protein is already targeted by an approved drug, or by drugs in clinical trials. We grouped variants identified by colocalisation into six regions, mapped to the genes: *LPA*, *IRF5*, *IRF4*, *IL20RB*, *NOS3*, and *APOE*. Variant-trait effects replicated in OFH for two regions: a variant (rs10455872) in the intron of *LPA*, showed causal effects for a group of cardiovascular conditions: abdominal aortic aneurysm (AAA), hypertension, peripheral arterial disease, and nonrheumatic valve disease, and a variant (rs3757387) upstream of *IRF5* was causal for the immune-mediated inflammatory diseases: hypothyroidism, RA, and UC.

The remaining four regions consisted of: a variant (rs9392504) downstream of *IRF4*, causal for hyperthyroidism, hypothyroidism, and RA; a variant (rs3918226) in *NOS3* causal for CAD, chronic kidney disease (CKD), hypertension, and stroke; and a well-known missense variant (rs429358) in *APOE*, which showed causal effects for cholelithiasis, chronic obstructive pulmonary disease (COPD), T2D, and OA. Finally, a variant (rs1394094) in the intron of *STAG1*, found to be a *cis*-pQTL and eQTL for *IL20RB*, causal for carpal tunnel syndrome, hypertension, T2D, and OA.

### Highlighting potential adverse effects

In cases where a genetic variant increases risk of some conditions but decreases risk of others (known as antagonistic pleiotropy), drugs targeting the related protein could have unintended adverse effects. We explored regions in which the colocalising variant(s) showed discordant directions of effect on conditions, and the implicated protein is already targeted by an approved drug, or by drugs in clinical trials. We identified eight regions where this was the case; for two of these regions, variant-condition associations replicated in OFH. One allele, rs11205303-C, increased risk of CAD and breast cancer, but decreased risk of carpal tunnel syndrome. Despite being a missense variant in *MTMR11*, the variant is a *cis*-pQTL, eQTL, and is predicted to overlap with the enhancer region for *SV2A*; we labelled *SV2A* as the likely effector gene, though eQTL and *cis*-pQTL associations with other genes and proteins suggests that the variant may be influencing multiple pathways. The second region in this category contained a missense variant in *CPS1*, with the A-allele increasing risk of CKD and OA but decreasing risk of cholelithiasis. Other regions included variants mapping to the genes: *GCKR*, *FAP*, *TDGF1*, *SLC39A8*, *SH2B3*, and *GIPR*. Full results can be found in Supplementary Tables 4 and 6.

We also explored regions in which the colocalising variant(s) showed discordant directions of effect on conditions, but for which the implicated protein is not currently clinically targeted, though it is considered druggable. We found regions in this category, with four regions implicating the genes *ZC3HC1*, *L3MBTL3*, *BACH2*, and *CTSB* replicating variant-condition associations in OFH. Other regions included a missense variant in *RREB1*, a variant upstream of *KLF14*, and a complex region implicating *BTN3A2* and other genes. Full results can be found in Supplementary Table 6.

Four regions showed a higher level of biological complexity, containing more than two variants and implicating multiple genes. Five moderately correlated (R^2^>0.3) variants on chromosome 8 mapped to the genes *MSRA* and *CTSB*, with additional genes in the locus (Supplementary Figure 6). The variants showed causal effects in a concordant direction for a variety of conditions: COPD, hypertension, T2D, OA, and sleep apnoea, with an opposing effect on hypothyroidism.

Colocalisation identified three variants in the *CDKN2A/B* locus, also known as the 9p21 locus, causal for AAA, CAD, ischaemic stroke and TIA, stroke (all), hypertension, peripheral arterial disease, glaucoma, and in the opposite direction, breast cancer. Supplementary Figure 7 illustrates the LD patterns of these three variants.

A region containing 11 variants was on chromosome 6, spanning ∼2.5Mb. Variants showed concordant effects across immune-mediated conditions, but opposite effects on psychiatric conditions and breast cancer. Supplementary Figure 8 illustrates the complex LD patterns between these 11 variants. Though we excluded the HLA region due to its complex LD, remaining signals adjacent to the HLA region could still reflect HLA-related pathways.

The region containing the most variants was on chromosome 12, spanning ∼1.5Mb, including the genes *CUX2*, *SH2B3*, *ATXN2*, *ACAD10*, *NAA25*, *HECTD4*, and *PTPN11*. Supplementary Figure 9 illustrates the LD patterns between the variants identified in this region. Sixteen variants implicated 10 conditions crossing different disease domains (see Supplementary Table 4). Variants had concordant directions of effect on eight of the conditions, but opposite directions of effect on two cancer phenotypes, breast cancer and colorectal cancer.

### Uncovering shared genetic mechanisms

Finally, we note cases where the genetic variant(s) influence proteins that are not currently thought to be druggable. Though these cannot inform potential treatments in the near future, they can still give interesting insights into shared biological mechanisms in MLTC. Details of these regions can be found in Supplementary Table 4.

## Discussion

Using large-scale human genetics data and genetic colocalisation, we have characterised shared genetic architecture across 72 long-term conditions, providing insights into the biological basis of MLTC. These findings have several clinical implications, including the identification of potential therapeutic targets, opportunities for drug repurposing, and cases where therapeutic interventions may have unintended adverse effects.

Pairwise colocalisation analysis identified 1,181 loci with evidence of a shared causal genetic variant across 434 long-term condition-pairs. For 281 pairs, specific causal variants were identified, totalling 578 variants. These results provide a comprehensive resource for exploring pleiotropy and shared causal mechanisms in MLTC. We applied a set of criteria to highlight results for discussion, but considerable nuance is required to interpret each specific locus and MLTC combination. For instance, some regions are large and may include more than one causal variant (such as the *SH2B3* locus) or multiple independently causal genes (such as the *CTSB* locus). Further, whilst a shared causal variant for MLTC implies a shared biological pathway (and possible intervention), we cannot always rule out horizontal pleiotropy – where a single variant impacts the conditions via independent pathways.

We prioritised genetic variants causal for three or more LTCs, to focus on more highly pleiotropic genetic signals that may be more relevant to MLTC. Variants influencing multiple conditions may represent shared biological mechanisms and could therefore have relevance for therapeutic target prioritisation, where a single target may potentially influence several conditions. We identified 34 independent genetic regions containing one or more variant found to colocalise between more than one LTC pair (three or more individual LTCs). Proteins associated with eight of these regions present opportunities as novel therapeutic targets. For example, a region on chromosome 2 containing two variants, where the variant-trait effects replicated in OFH. The variants (rs9306894-G and rs2289081-C) are in high LD (R^2^=0.90) and are located in the 3′ UTR and downstream of *GDF7*, respectively. Both variants are eQTLs for *GDF7* (listed alleles associated with higher expression), and as they are in an enhancer region for *GDF7*, may causally increase transcription of the gene. In our analysis, the alleles increased risk of diverticular disease, GORD, female genital prolapse, and prostate cancer. This is in line with previous GWAS findings of associations between rs9306894-G and increased risk of pelvic organ prolapse [30] and prostate cancer [31], and between rs2289081-C and increased risk of diverticular disease [32] and prostate cancer [33]. *GDF7*, growth differentiation factor 7, encodes the protein GDF7, also known as BMP12 (bone morphogenetic protein 12), a member of the transforming growth factor-beta (TGF-beta) superfamily of proteins. GDF7 has an established role in connective tissue homeostasis, including tendon and ligament development and repair [34]. Associations with diverticular disease, GORD, and female genital prolapse might reflect a shared connective tissue aetiology, given that all three conditions involve structural weakness of supporting connective tissues. The link to prostate cancer is less clear, though BMP signalling has been previously implicated in prostate cancer [35]. GDF7 is not currently targeted by any approved drugs or by drugs in clinical trials, but evidence from the Open Targets platform [13] predicts with medium confidence that it is druggable and tractable by antibody therapeutics.

We explored potential opportunities for drug repurposing – where existing approved drugs, or drugs in clinical trials, could be used for the treatment of additional conditions. Therapeutic evidence for each protein is summarised in Supplementary Table 7. We identified six regions across our analysis, and here we highlight two regions where we replicated the variant-trait effects identified in OFH. One variant, rs3757387, upstream of the gene *IRF5*, was implicated in a cluster of immune-mediated inflammatory diseases: hypothyroidism, RA, and UC. The variant has been previously associated with hypothyroidism [36], RA [37], and UC [38], along with the autoimmune conditions, systemic lupus erythematosus [39] and Sjögren’s syndrome [40], and a measure of kidney function, estimated glomerular filtration rate [41]. It is thought that rs3757387 lies in the *IRF5* promotor region, and influences methylation levels of a nearby CpG site, which then alters *IRF5* expression [41]. Interferon regulatory factor 5 (*IRF5*) encodes a transcription factor with a central role in immune responses and is implicated in many autoimmune and inflammatory diseases [42]. Traditionally, transcription factors are considered not druggable due to their lack of a conventional active binding site. However, technological advancements mean that proteins previously thought to be undruggable, can now be targeted. In the case of IRF5, companies are developing novel approaches to target the protein, including selective small-molecule inhibitors and an oral degrader, which have demonstrated promising preclinical activity in models of autoimmune and inflammatory disease [43,44]. IRF5 is also predicted tractable by PROTAC, another emerging drug modality, highlighting the future potential to target IRF5.

Colocalisation analysis found a variant in the intron of *LPA*, rs10455872, to be causal for a group of cardiovascular conditions: AAA, hypertension, peripheral arterial disease, and nonrheumatic valve disease. *LPA* encodes apolipoprotein(a), a key component of lipoprotein(a), a major risk factor for cardiovascular disease. The variant is associated with higher levels of *LPA* mRNA and protein levels of apolipoprotein(a) [45], and has previously been reported in GWAS of individual cardiovascular conditions [46]. Drugs targeting LPA for prevention of major adverse cardiac events (MI, stroke, CVD death, or coronary revascularization surgery) are in phase 3 clinical trials (e.g. Muvalaplin and Pelacarsen; see Supplementary Table 7) [47]. Our results provide evidence that targeting LPA may have broader cardiovascular benefits, supporting potential repurposing.

We highlight 15 cases where genetic variants show discordant effects across conditions – increasing risk of some conditions but decreasing risk of others. These cases are important for drug development, as drugs targeting the implicated protein could have risks of unintended adverse effects. For eight identified regions, a drug exists, either approved or in clinical trials, that targets the related protein; for a further seven regions, no drugs currently exist.

An allele in *GIPR*, rs10423928-A, decreased risk of obesity and sleep apnoea but increased risk of breast cancer. Previous studies have found rs10423928-A to be associated with impaired glucose- and GIP-stimulated insulin secretion, and a decrease in BMI, lean body mass, and waist circumference [48]. A *GIPR* missense variant in complete LD (R^2^=1, D′=1 in EUR), rs1800437-C, has been associated with increased risk of breast cancer [49]. This suggests that impaired *GIPR* function may be favourable for weight loss but could increase risk of breast cancer.

*GIPR* (gastric inhibitory polypeptide receptor) encodes the receptor for GIP (gastric inhibitory polypeptide, also known as glucose-dependent insulinotropic polypeptide), which is one of two incretin hormones along with glucagon-like peptide-1 (GLP-1) that are released after eating, responding to elevated blood glucose by stimulating insulin secretion [50]. Tirzepatide, a drug recently approved for weight management and type 2 diabetes treatment, is a dual GIPR and GLP-1R agonist. However, there is evidence that supports both GIPR agonism and antagonism leading to a reduction in body weight [51]. One of the first studies investigating how GIP could regulate body weight found that GIPR knockout mice were protected from high-fat diet-induced obesity [52]. Our findings are consistent with these observations, suggesting that genetically reduced GIPR signalling, approximating pharmacological GIPR antagonism, may be beneficial for weight loss. Though tirzepatide is the only medication currently approved with this target, there are others in clinical trials, including AMG 133, a GIPR antagonist and GLP-1R agonist being investigated for clinical weight loss [53]. The potential increased risk of breast cancer associated with reduced GIPR signalling should be considered in the case of medications that reduce GIPR signalling. Given that evidence supports both GIPR agonism and antagonism reducing body weight, increased understanding of GIPR biology is needed, especially on the interaction between GIPR and GLP-1R.

For several regions, including those implicating proteins SH2B3, BTN3A2, and CDKN2B (though not considered druggable), we see genetic variants that decrease risk of multiple conditions but increase risk of cancer phenotypes. For instance, a missense variant in *SH2B3* (rs3184504-C) was associated with decreased risk of cardiovascular conditions – CAD, heart failure, hypertension, and thromboembolic diseases, musculoskeletal conditions – gout, OA, and RA, and hypothyroidism, but increased risk of breast cancer. This is in line with previous research into the genetics of ageing; the same missense variant has been found to be associated with lifespan, and it has been hypothesised that the variant is an example of antagonistic pleiotropy – the theory that some genetic mutations that are beneficial in early life become harmful in later life [54,55]. Here, this reflects a trade-off between chronic disease and cancer. It is important to note that for these three regions, whilst we have selected a single variant linked to a single protein, regions are large and contain multiple causal variants, which could be associated with numerous genes/proteins. Further work is needed to disentangle the mechanisms linking these regions to disease.

A limitation of our analysis is that it is restricted to the set of conditions included; the shared causal mechanisms identified could implicate additional conditions not considered in our analysis. For example, rs429358 in *APOE* was identified as a shared causal variant for multiple conditions; this is a well-known variant involved in risk of Alzheimer’s disease (*APOE*-ε4) [56], a condition we did not include in this study due to the phenotype not passing a heritability threshold implemented during condition selection previously [4]. This could affect the suggested clinical translation; for the *APOE* region, colocalisation found rs429358-T to increase risk of cholelithiasis, COPD, T2D, and OA, and so the regions is categorised as a repurposing opportunity, as it shows concordant effects on all LTCs – if Alzheimer’s disease were included as a condition, rs429358-C increases risk [57], and so the region would become one associated with possible adverse effects. Future work could include a broader set of conditions.

We note that the *coloc.abf* framework assumes at most a single causal variant per trait per genomic region [10], meaning that our analysis may have missed additional shared causal variants in regions where multiple signals exist. Methods exist that allow for multiple causal variants, such as *coloc.susie*, but they require high-quality LD reference data compatible with the GWAS data used and are known to be very sensitive to any mismatches [58]. Additionally, the authors of SuSiE caution against using meta-analysed data, because SNPs that are not measured in all studies included in a meta-analysis can affect fine mapping [59]. We could not consistently implement *coloc.susie* across all regions for all condition pairs, limiting robust application. To mitigate this limitation, we chose a small window size (±250kb centred on the lead variant) for regions given to *coloc.abf*, to reduce the likelihood of including multiple independent signals within a single region. Additionally, the main objective of this study was to identify shared genetic mechanisms across multiple conditions and evaluate their therapeutic potential, rather than to comprehensively resolve variant-level causal genetic architecture. Downstream analyses therefore focused on prioritising candidate genes and proteins implicated by the colocalisation findings.

Whilst a strength of this study is that we used OFH data to replicate variant-LTC associations for the variants found in colocalisation analyses, future work should include replication of analyses in separate datasets to assess the robustness of findings, and experimental follow-up of promising results.

## Conclusions

In conclusion, we identified shared genetic mechanisms across multiple long-term conditions and translated these findings into biologically and clinically meaningful insights. Starting from GWAS association signals, we integrated genetic colocalisation with downstream biological and therapeutic analyses to progress from shared genetic associations to identifying candidate genes, proteins, and biological pathways, enabling assessment of their translational relevance. This work provides a translational framework linking genetic discovery with potential clinical application and highlights promising avenues for future mechanistic investigation.

## Data sharing statement

Most data generated during this study are included in this published article and its supplementary information files. We cannot make individual-level data available. Researchers can apply to UK Biobank (https://www.ukbiobank.ac.uk/enable-your-research/)and Our Future Health (https://research.ourfuturehealth.org.uk/apply-to-access-the-data/). We have made our diagnostic code lists, scripts, and results available on our GitHub (https://github.com/GEMINI-multimorbidity/).

## Declaration of interests

JB is a part-time employee of Novo Nordisk Research Centre Oxford, limited, unrelated to this work. TMF has consulted for several pharmaceutical companies. All other authors have no disclosures to declare.

## Funding

This work was supported by the UK Medical Research Council [grant number MR/W014548/1]. TMF, SEL, CF, KB, JAHM, and DT are supported by the National Institute for Health and Care Research (NIHR) Exeter Biomedical Research Centre (BRC). This study was supported by the NIHR Exeter BRC, the NIHR Leicester BRC, the NIHR Oxford BRC, the NIHR South West Peninsula Applied Research Collaboration (PenARC) and the NIHR HRC in sustainable innovation.

TMF is additionally supported by the Swiss National Science Foundation Proposal 10.003.484: Genetic studies to identify causes and consequences of excess unhealthy weight, the Novo Nordisk Foundation NNF24OC0089256: Decoding the genetic and cellular basis of excess unhealthy weight, the EU-supported Innovative Medicines Initiative (IMI) funded project SOPHIA (Stratification of Obesity Phenotypes to Optimise Future Obesity Therapy), and the Fondation pour la Recherche sur le Diabète.

KB is partly funded by the NIHR Applied Research Collaboration South West Peninsula. JAHM is funded by an NIHR Advanced Fellowship (NIHR302270). CF is an NIHR Senior Investigator and is supported by the NIHR Health technology Research Centre (HRC) in Sustainable Innovation, the NIHR HRC coordinating centre England and the NIHR/ESRC/Alzheimer’s society Dementia Connect plus Sustainable Prevention, Innovation and Involvement Network. CV acknowledges research funding by a ‘Contratos para la intensificación de la actividad investigadora en el Sistema Nacional de Salud’ (INT23/00040) from the Spanish Ministry of Science and Innovation.

The views expressed are those of the authors and not necessarily those of the NIHR or the Department of Health and Social Care. The funders were not involved in the study design; in the collection, analysis, and interpretation of data; in the writing of the report; or in the decision to submit the paper for publication.

## Acknowledgements

This is a summary of independent research conducted at the National Institute for Health and Care Research (NIHR) Exeter Biomedical Research Centre (BRC) and by the NIHR Leicester BRC. The work was funded by the UK Medical Research Council (MRC). The interpretation and conclusions contained in this study are those of the authors and not necessarily those of the MRC, NIHR or the Department of Health and Social Care, nor of any other funders. This research has been conducted using the UK Biobank Resource, under application 14631. The authors would like to acknowledge and thank the participants and investigators of the UK Biobank and the FinnGen study. This study makes use of de-identified data held by Our Future Health (study ID OFHS250013). We would like to acknowledge all the research participants who have donated their data to the Our Future Health research programme. This work uses data that has been provided by patients and collected by the NHS as part of their care and support. The data are collated, maintained and quality assured by the National Disease Registration Service, which is part of NHS England. Access to the data was facilitated by the NHS England Data Access Request Service. The authors would like to acknowledge both past and present members of the GEMINI Consortium. The authors would like to acknowledge the use of the University of Exeter High-Performance Computing (HPC) facility in carrying out this work.

For the purpose of open access, the author has applied a Creative Commons Attribution (CC BY) license to any Author Accepted Manuscript version arising from this submission.

## Author Contributions

Funding acquisition: JD, CV, MM, LF, KB, CF, SEL, FD, JB, TMF, JAHM, and LCP. Conceptualisation: TMF, JAHM, and LCP. Project administration and supervision: TMF, JAHM, and LCP. Data curation: BV, RMW, LCP. Methodology: BV, RMW, FD, JB, TMF, JAHM, LCP. Resources: LCP. Formal analysis, investigation, and software: BV, RMW, LCP. Visualisation: BV. Writing - original draft: BV, RMW, TMF, JAHM, LCP. Writing - review and editing: BV, RMW, OM, JD, DT, RH, CV, MM, LF, KB, CF, SEL FD, JB, TMF, JAHM, LCP. All authors read and approved the final manuscript.

## Notes

### Author Declarations

The Northwest Multi-Centre Research Ethics Committee approved the collection and use of UK Biobank data for health-related research (Research Ethics Committee reference 11/NW/0382). Access to UK Biobank data was granted under UK Biobank approved application 14631. This study makes use of de-identified data held by Our Future Health (study ID: OFHS250013). Ethical approval for Our Future Health was granted by the East of England - Cambridge East Research Ethics Committee (Research Ethics Committee reference: 21/EE/0016).

