## Supplementary Information for "Identifying genetic variants causal for multiple long-term conditions: informing opportunities for intervention and prevention"

### Supplementary Figures

#### Supplementary Figure 1: global loci illustration

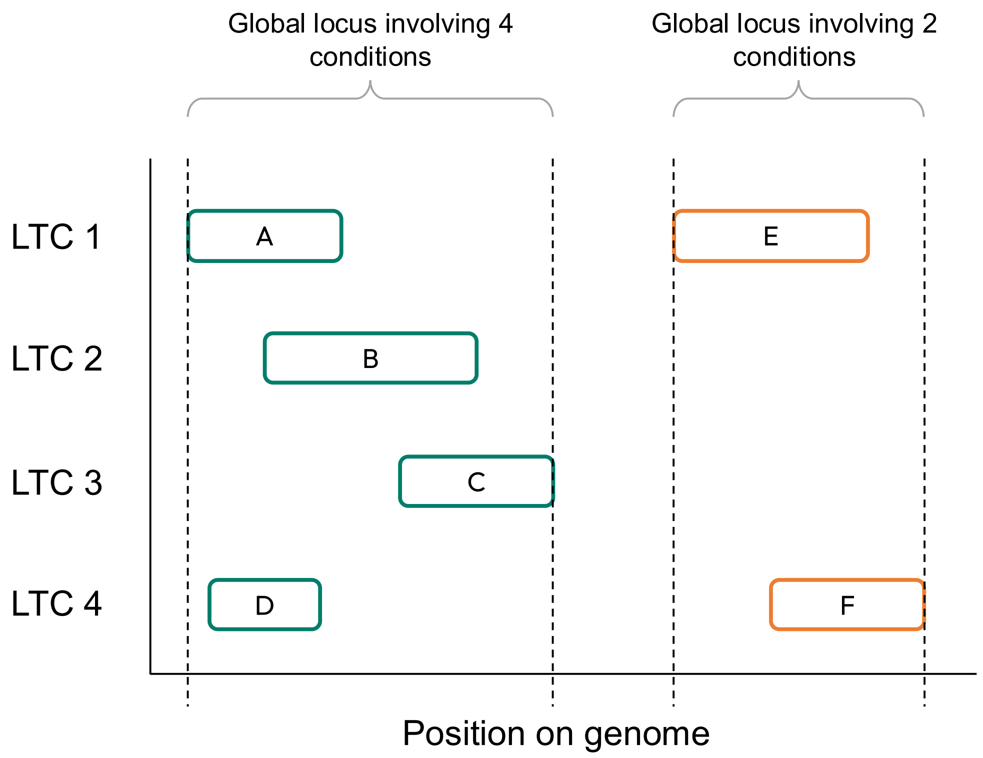

An illustration of how global loci would be defined across a set of four long-term conditions (LTCs). Locus A (for LTC1) physically overlaps with locus B (for LTC2), and locus D (for LTC4). Locus B also overlaps with locus C (for LTC3), so all four loci are grouped into one global locus.

### Supplementary Figure 2: number of genome-wide significant genetic variants per condition

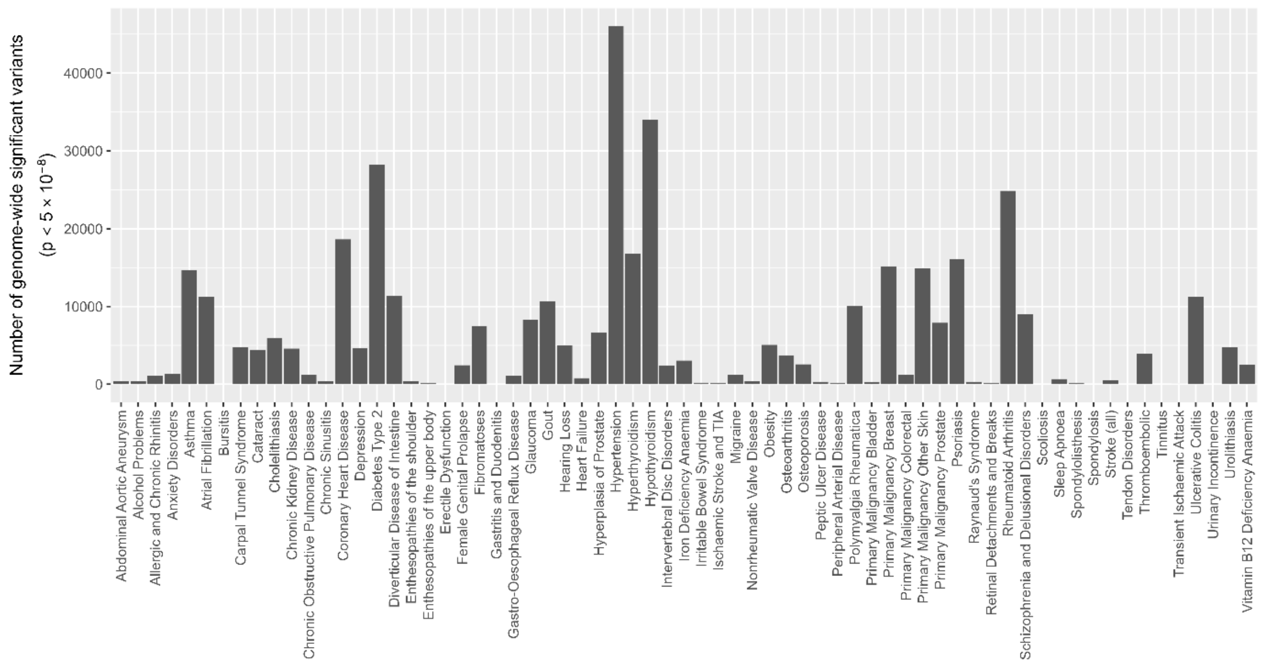

An illustration of the number of genome-wide genetic variants ($p<5\times10^{-8}$) for the 67 of 72 LTCs with at least one genome-wide significant variant.

### Supplementary Figure 3: proportion of genome-wide significant variants per condition associated with other conditions

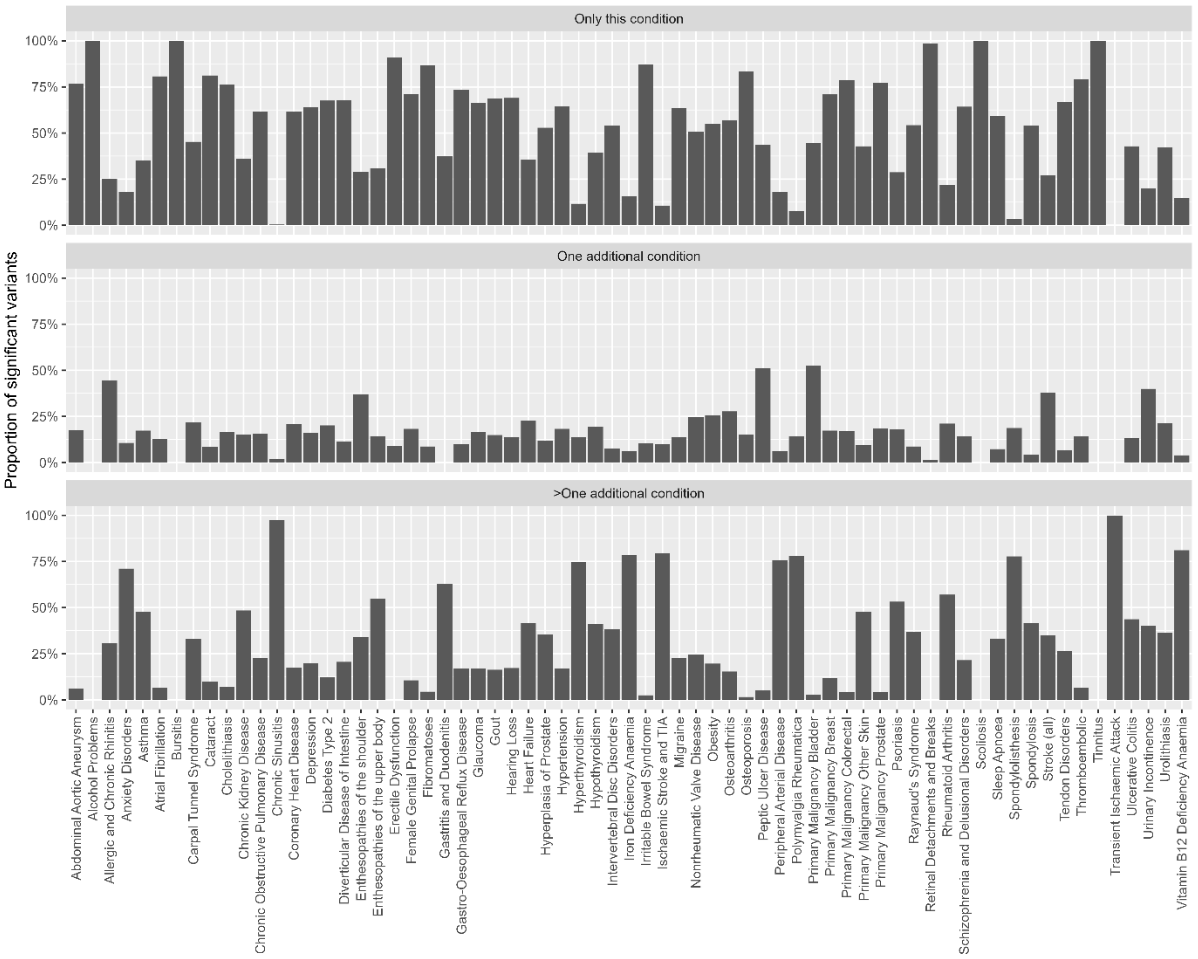

An illustration of the proportion of genome-wide significant genetic variants per condition that are associated with just that condition, one additional condition, or more than one additional condition.

### Supplementary Figure 4: number of genetic loci per condition

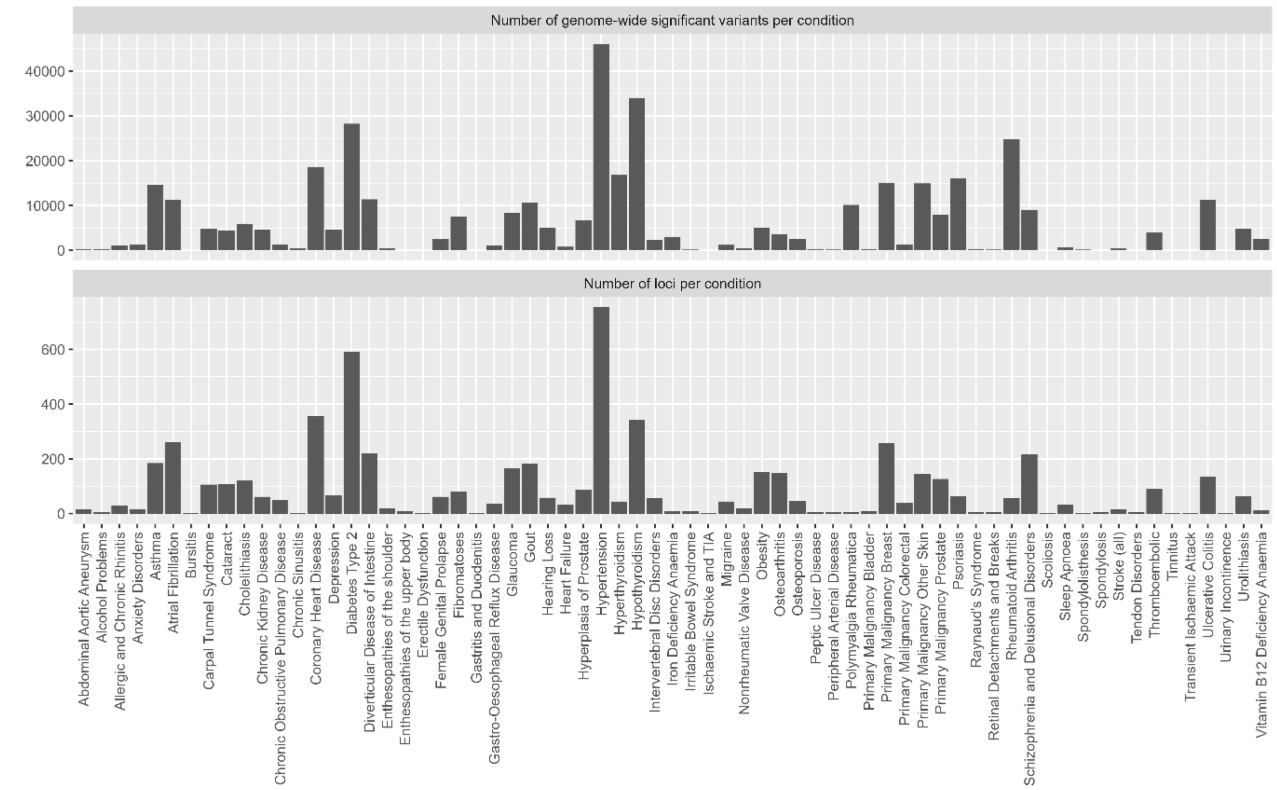

An illustration of the number of genome-wide significant variants for each of the 67 LTCs with at least one, and how this maps to the number of genetic loci per condition.

### Supplementary Figure 5: global genetic loci across 67 long-term conditions

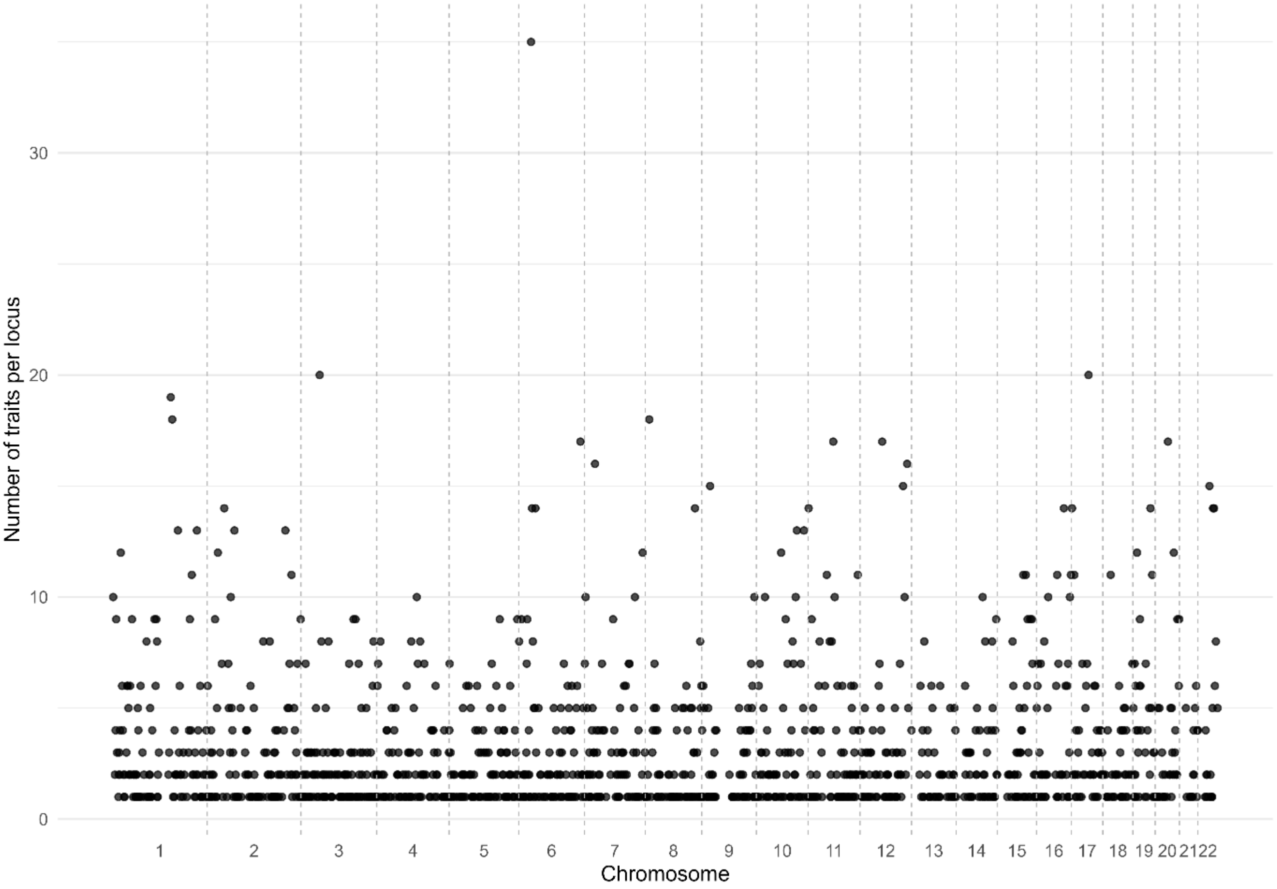

An illustration showing the areas of the genome containing genetic loci shared for multiple conditions. Each point denotes a genetic locus, the y-axis denotes the number of conditions that have significant genetic signals in that genetic locus, and the x-axis denotes the position of the locus on the genome.

Supplementary Figure 6: Linkage disequilibrium patterns of the variants in the *MSRA*/*CTSB* region

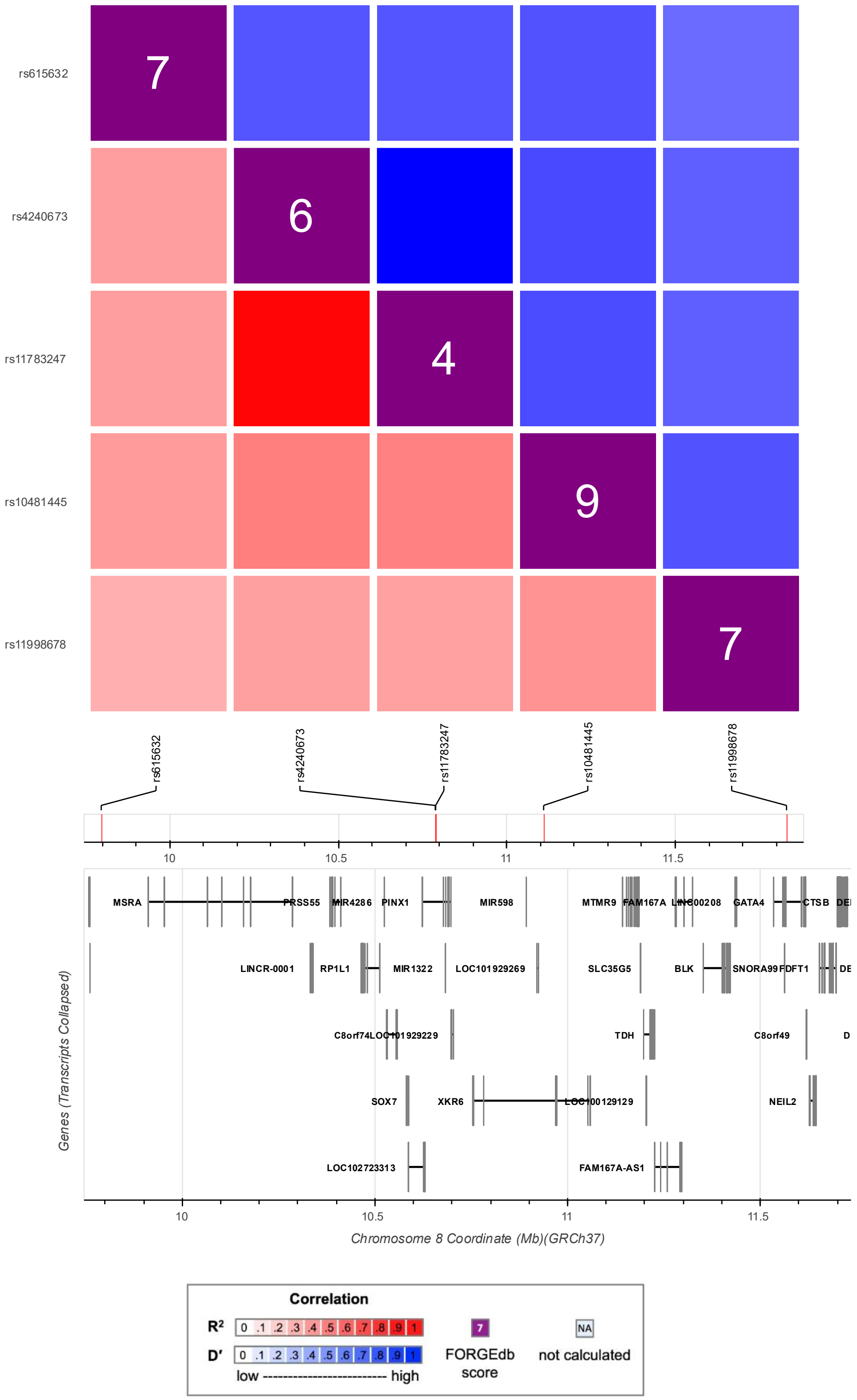

A heatmap generated using the LDmatrix tool (1) showing the linkage disequilibrium patterns for the five genetic variants in the *MSRA/CTSB* region identified by colocalisation.

### Supplementary Figure 7: Linkage disequilibrium patterns of the variants in the *CDKN2A/B* region

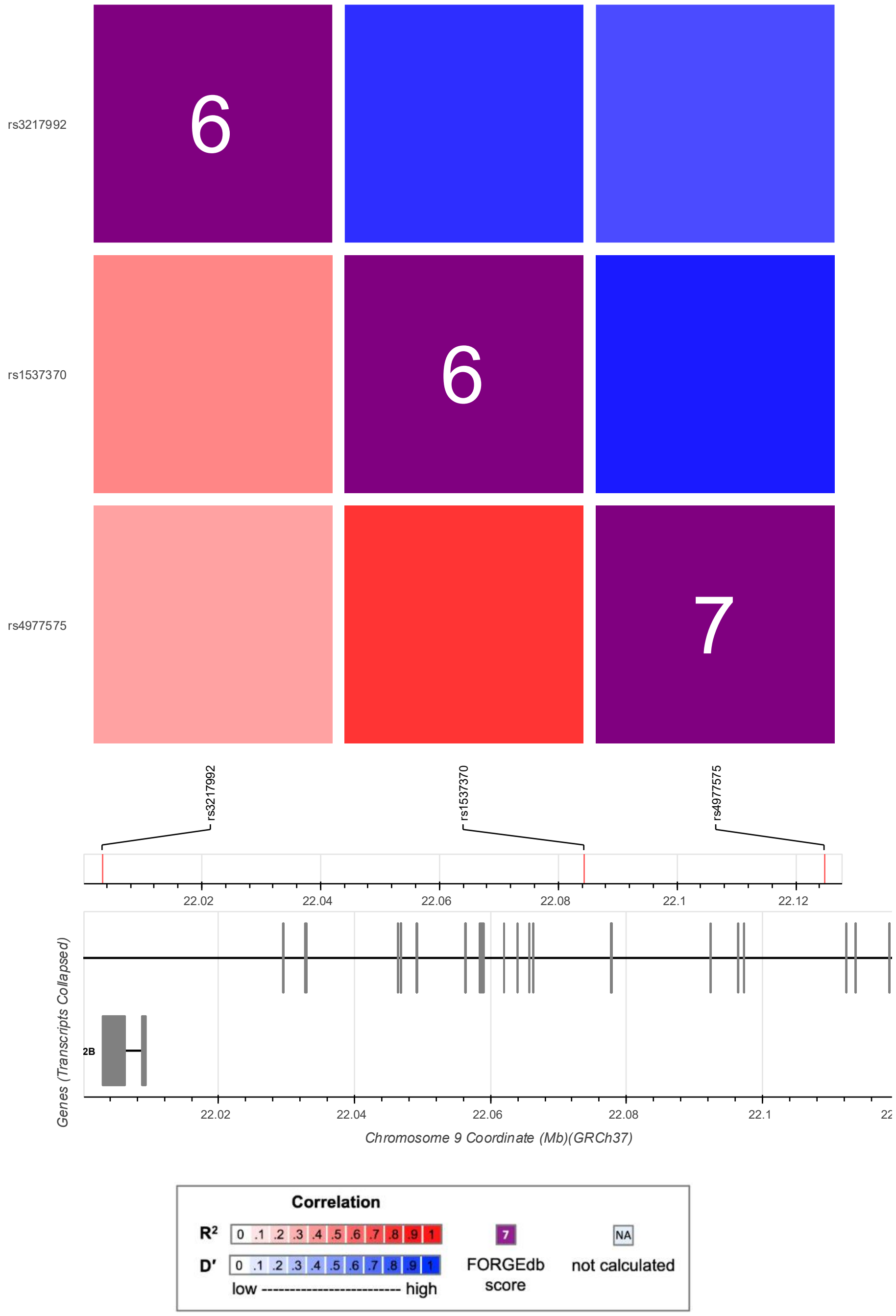

A heatmap generated using the LDmatrix tool (1) showing the linkage disequilibrium patterns for the three genetic variants in the *CDKN2A/B* region identified by colocalisation.

Supplementary Figure 8: Linkage disequilibrium patterns of the variants in the *BTN3A2* region

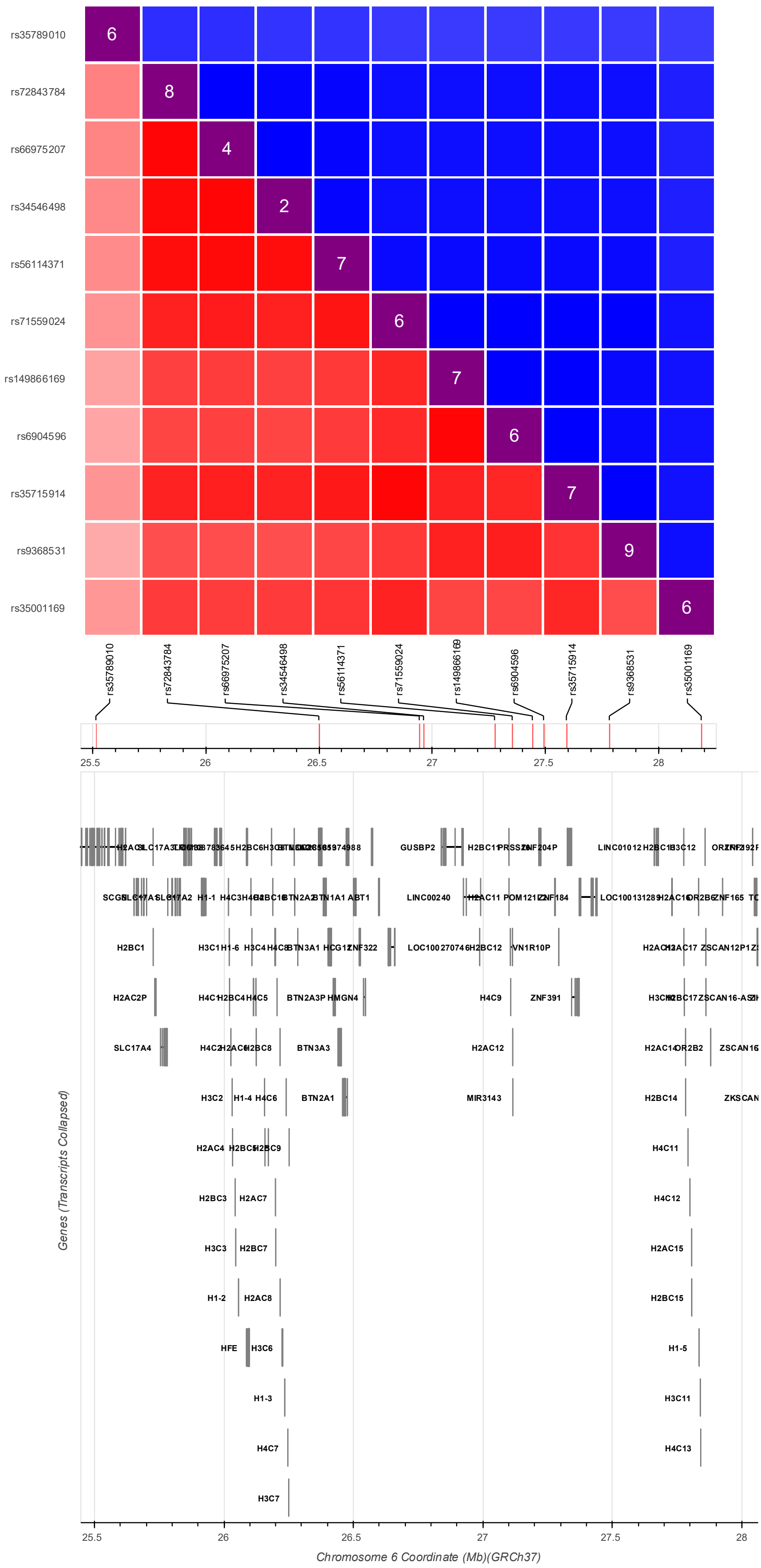

rs35789010
rs72843784
rs66975207
rs34546498
rs56114371
rs71559024
rs149866169
rs6904596
rs35715914
rs9368531
rs35001169
6
8
4
2
7
6
7
6
7
9
6
25.5
26
26.5
27
27.5
28
Genes (Transcripts Collapsed)
Chromosome 6 Coordinate (Mb)(GRCh37)
GUSBP2
LINC01012
SCGN
H1-1
BTN2A2
BTN1A1
ABT1
LINC00240
ZNF184
ZNF165
H2BC1
BTN3A1
ZNF322
VN1R10P
H2AC2P
H4C1
H2BC4
H4C5
BTN2A3P
HMGN4
H4C9
ZNF391
SLC17A4
H4C2
H2BC8
BTN3A3
H2AC12
OR2B2
H3C2
H1-4
H4C6
BTN2A1
MIR3143
H2BC14
ZKSCAN
H2AC4
H4C11
H2BC3
H2AC7
H4C12
H3C3
H2BC7
H2AC15
H1-2
H2AC8
H2BC15
HFE
H3C6
H1-5
H1-3
H3C11
H4C7
H4C13
H3C7

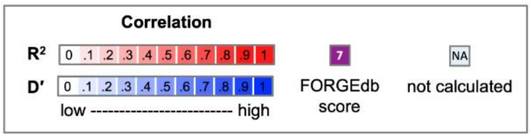

A heatmap generated using the LDmatrix tool (1) showing the linkage disequilibrium patterns for the 11 genetic variants in the *BTN3A2* region identified by colocalisation.

### Supplementary Figure 9: Linkage disequilibrium patterns of the variants in the *SH2B3* region

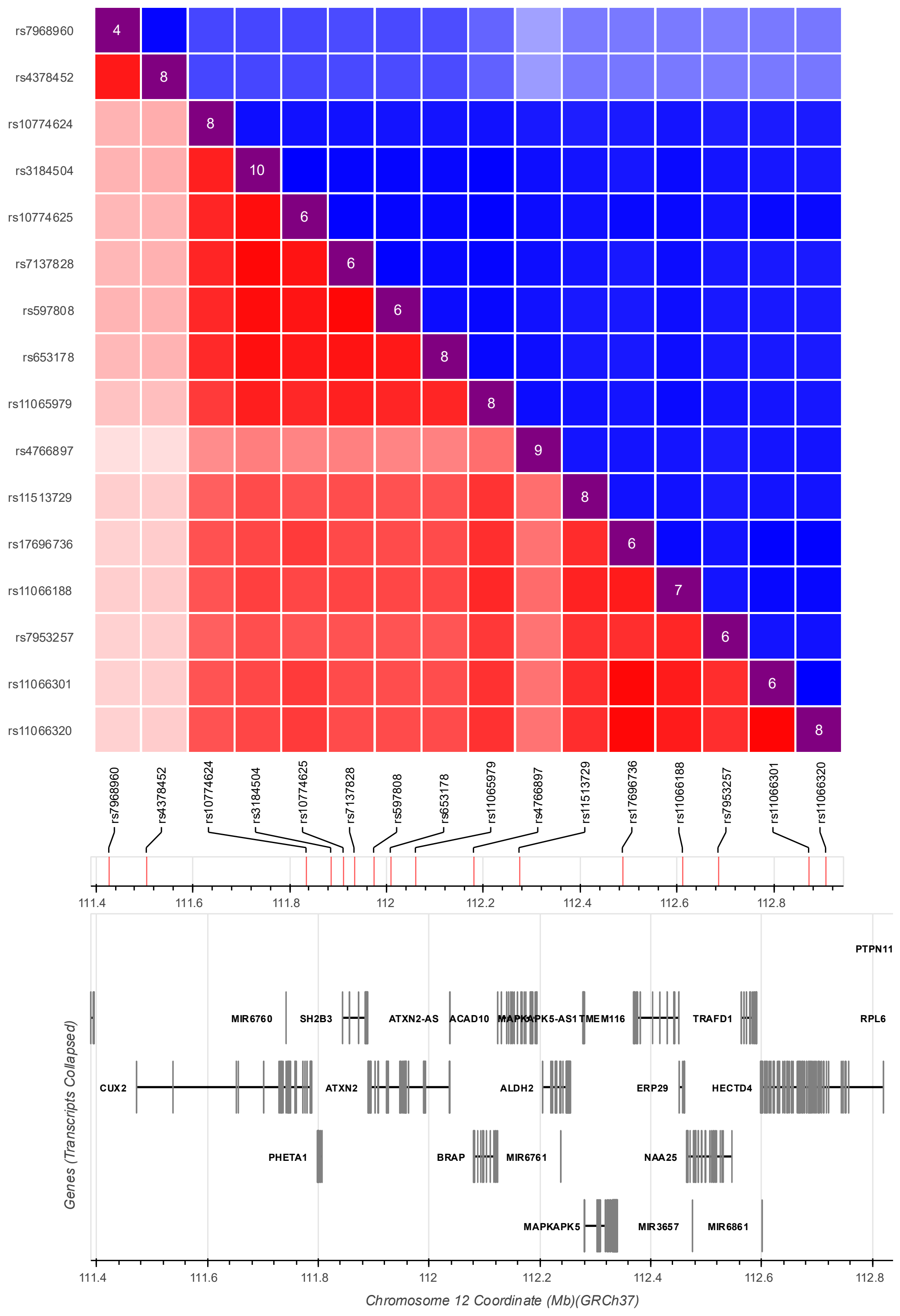

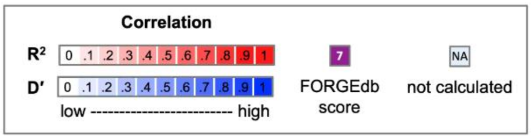

A heatmap generated using the LDmatrix tool (1) showing the linkage disequilibrium patterns for the 16 genetic variants in the *SH2B3* region identified by colocalisation.
